# The Impact of Conventional Stroke Risk Factors on Early and Late Onset Ischemic Stroke: a Mendelian Randomization Study

**DOI:** 10.1101/2024.05.31.24308308

**Authors:** Kevin TK Nguyen, Huichun Xu, Brady Gaynor, Sally N. Adebamowo, Patrick F. McArdle, Tim O’Connor, Bradford Worrall, Rainer Malik, Giorgio B. Boncoraglio, Ramin Zand, Steven J. Kittner, Braxton D. Mitchell the Stroke Genetic Network and Early-Onset Stroke Consortia.

## Abstract

**Objective:** Although stroke incidence is decreasing in older ages, it is increasing in young adults. While these divergent trends in stroke incidence are at least partially attributable to diverging prevalence trends in stoke risk factors, age-dependent differences in the impact of stroke risk factors on stroke may also contribute. To address this issue, we utilized Mendelian Randomization (MR) to assess differences in the association of stroke risk factors between early onset ischemic stroke (EOS) and late onset ischemic stroke (LOS).

**Methods:** We employed a two-sample MR design with inverse variance weighting as the primary method of analysis. Using large publicly available genome-wide association summary results, we calculated MR estimates for conventional stroke risk factors (body mass index, total, HDL-and LDL-cholesterol, triglycerides, type 2 diabetes, systolic and diastolic blood pressure, and smoking) in EOS cases (onset 18-59 years, n = 6,728) and controls from the Early Onset Stroke Consortium and in LOS cases (onset ≥ 60 years, n = 9,272) and controls from the Stroke Genetics Network. We then compared odds ratios between EOS and LOS, stratified by TOAST subtypes, to determine if any differences observed between effect sizes could be attributed to differences in the distribution of stroke subtypes.

**Results:** EOS was significantly associated with all risk factors except for total cholesterol levels, and LOS was associated with all risk factors except for triglyceride and total cholesterol levels. The associations of BMI, DBP, SBP, and HDL-cholesterol were significantly stronger in EOS than LOS (all p < 0.004). The differential distribution of stroke subtypes could not explain the difference in effect size observed between EOS and LOS.

**Conclusion:** These results suggest that interventions targeted at lowering body mass index and blood pressure may be particularly important for reducing stroke risk in young adults.

## Introduction

Stroke is one of the leading causes of death and disability, with more than 795,000 new and recurrent cases annually in the United States as of 2023.^1^ Despite its high incidence, stroke is largely a preventable disorder. The Global Burden of Disease Study estimated that 91% of stroke burden, measured as disability-adjusted life years, can be attributable to modifiable risk factors and 72% of stroke burden is attributable to clusters of metabolic risk factors, namely hypertension, obesity, hyperglycemia, hyperlipidemia, and renal dysfunction.^2,3^ While most strokes occur in individuals over the age of 50 years, approximately 10% of strokes occur in younger adults aged 18-50 years.^4^ Since the year 2000, incidence rates of ischemic stroke in high-income countries have been declining among older individuals but have been increasing in individuals younger than age 55.^5^ While these diverging trends in stroke incidence can be at least partially attributable to diverging patterns of stroke risk factors between younger and older adults, it is also possible that the impact of stroke risk factors differs between younger and older individuals.

To evaluate the impact of modifiable risk factors on ischemic stroke at different ages, we employed Mendelian randomization^6^ (MR) to compare causal associations by age of stroke onset and across stroke subtypes. MR uses genetically predicted levels of traits to serve as proxies. Since alleles are randomly assigned at conception, these genetic proxies are generally independent of the risk factor-outcome relationship and thus not easily subject to reverse causality or confounding factors as seen in observational studies. We hypothesize that the contributions of five modifiable ischemic stroke risk factors (blood pressure, body mass index, type 2 diabetes, hyperlipidemia, and smoking) differ between early and late onset ischemic stroke (EOS and LOS) and that these differences can be explained by the differential distribution of stroke subtypes. Using Mendelian randomization, we estimated the causal effects of these risk factors and compared these estimates between early (age of stroke onset < age 60 yrs.) and late (age of stroke onset ≥ 60 yrs.) We further considered whether any differences observed between effect sizes could be attributed to differences in the distribution of stroke subtypes.

## Methods

### Study design

We employed a 2-sample MR design using IVW as our primary method of analysis to assess the causal estimated of conventional stroke risk factors on early and late onset ischemic stroke.

### Study sample for Primary Outcome

This study utilizes stroke cases and controls assembled from two large GWAS consortia: The Early Onset Stroke Consortium^7^ (EOSC) and the Stroke Genetics Network^8^ (SiGN). Stroke cases in these Consortia underwent brain imaging at each site to exclude diagnoses other than ischemic stroke and to assist with subtype classification. Additional screening was performed in some, but not all, studies to exclude cases believed to be due to a known monogenic cause (e.g., sickle cell disease) or to a known non-genetic cause (e.g., drug use, complications of procedures). Ischemic stroke subtyping was performed using the TOAST criteria^9^ in most, but not all, sites.

Consistent with criteria used in the EOSC, we defined early onset stroke for these analyses as cases with stroke onset 18-59 years, and late-onset stroke as those with age at first stroke 60 years or older. Subjects included in this report are restricted to a subset of 6,728 early-onset cases (and 33,764 controls) and 9,272 late-onset stroke cases (and 25,124 controls) who are of European ancestry and for whom individual-level genotypes were available (**Table S1**). The genotype data from stroke cases and controls were based on hg38 and imputed using the TOPMed reference panel on the University of Michigan Imputation Server.^10^

### Exposure Genetic instrument selection

We obtained summary genetic association results from large publicly GWAS available from the GWAS catalog^11^ (https://www.ebi.ac.uk/gwas/) for 11 stroke risk factors: body mass index (BMI), systolic blood pressure (SBP), diastolic blood pressure (DBP), total cholesterol (TCHOL), LDL cholesterol (LDL), HDL cholesterol (HDL), triglycerides (TG), type 2 diabetes, type 2 diabetes adjusted for BMI, cigarettes per day, and smoking initiation. Sample sizes for the genome-wide association analyses of each trait ranged from 339,224 to 1,232,091 (**Table S2**). We identified variants from GWAS that were associated with each risk factor at genome-wide significance (p < 5 × 10^−8^) and selected the most significant variant at each associated locus by removing SNPs in linkage disequilibrium with the lead SNP using the clumping procedure in PLINK with parameters clump-kb = 10,000 and clump-r^2^ > 0.001. We assessed the strength of each SNP by calculating its F-statistic, which is a function of the proportion of the variance explained by the genetic instrument, and the sample size.^11^ The total number of SNPs obtained from each GWAS, the number of SNPs pruned at each filtering step, and the corresponding F-statistic are provided in **Table S3**.

### Mendelian Randomization Assumptions

To minimize the potential for bias in making causal inferences, SNPs were selected to adhere to three assumptions of valid instrumental variables (**Figure 2**):^12^ (1) the SNPs used must be strongly associated with the exposure; (2) the SNPs must not be associated with measured and unmeasured confounders; and (3) the SNPs affect the outcome only through the effects of the exposure (e.g., no horizontal pleiotropy.) We used the F-statistic as a measure of the strength of the SNP-exposure association (Assumption 1) and performed sensitivity analyses (see below) to assess violations of Assumptions 2 and 3.

**Figure 1:**
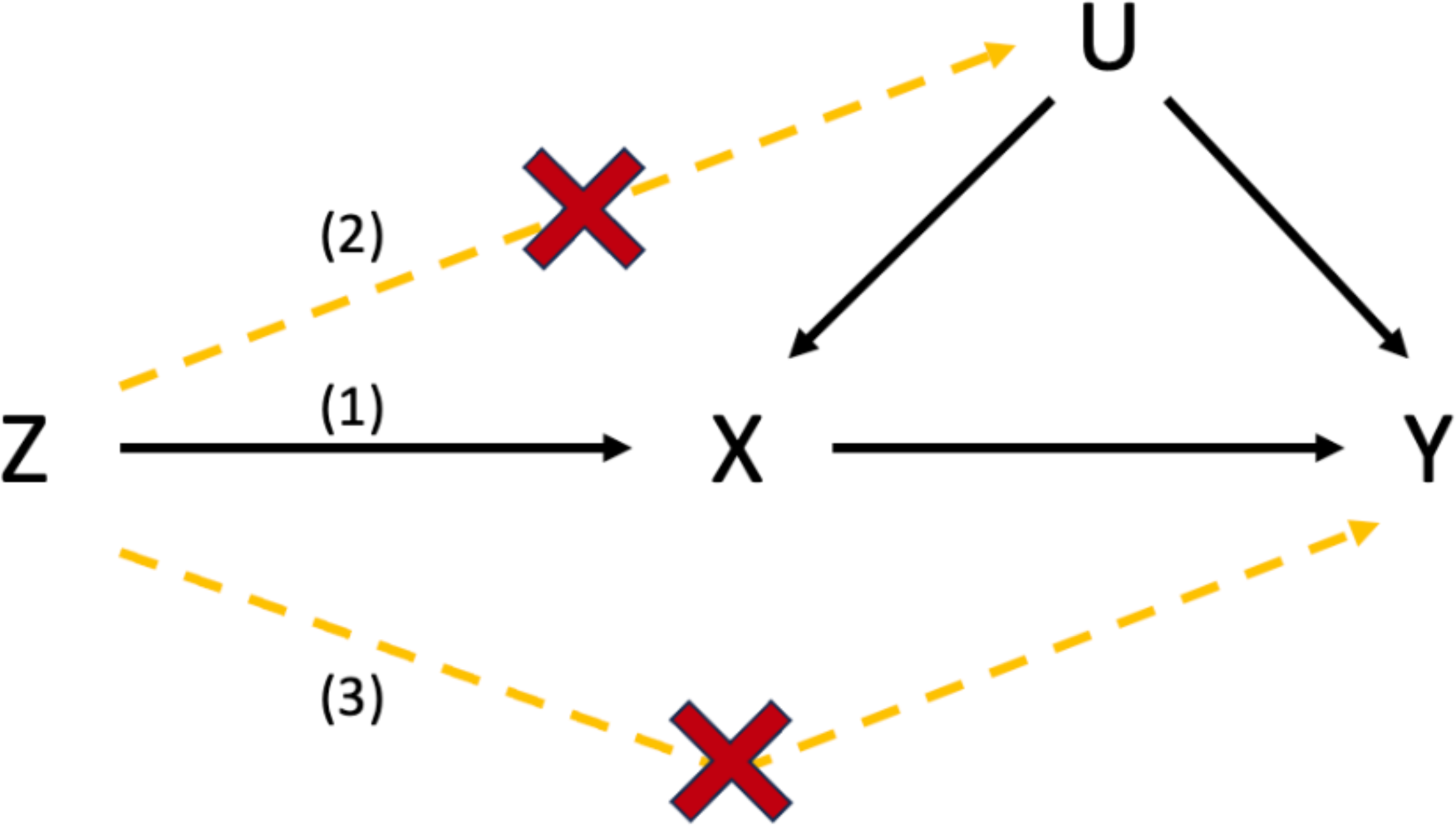
The Three MR Assumptions, where Z is the IV associated with the exposure, X is the exposure, Y is the outcome, and U is confounder. 1) Relevance Assumption: The IV is strongly associated with the exposure of interest. 2) Independence assumption: there are no confounders of the association between the IVs and the outcome, and 3) exclusion restriction assumption: the IV is not related to the outcome other than via the exposure

**Figure 2:**
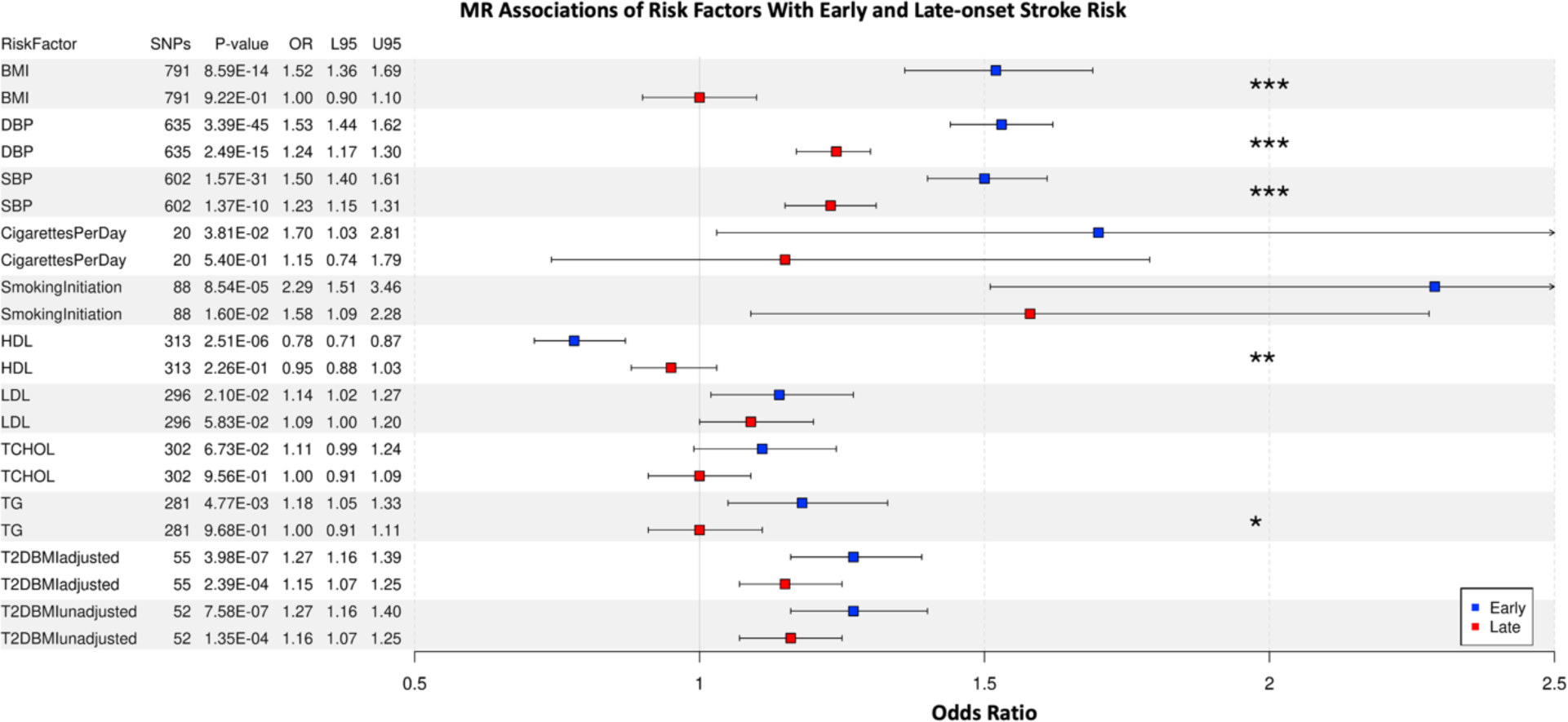
Odds ratio and 95% confidence interval for association of 11 stroke risk factors with EOS (blue) and LOS (red) for all ischemic strokes. *** P < 0.00075; ** P < 0.0075; * P < 0.05 for the heterogeneity test between EOS and LOS

### Statistical Analyses

Following clumping and F-statistic filtering, GWAS summary statistics were pulled for the associated SNPs to create the eleven exposure genetic risk scores. The SNP-outcome (stroke) associations were obtained from genetic association analyses performed on the early and late onset stroke datasets from the EOSC and SiGN. The SNP-outcome associations were calculated in PLINK2^13^ (PLINK v2.00a3.3LM) software for all ischemic stroke and for the TOAST subtypes using logistic regression, controlling for genetic ancestry with principal components 1 through 10 and sex.

We used the random-effects inverse-variance weighted^14^ (IVW) as the primary method for computing causal estimates of the association of each exposure (risk factor) with stroke. This approach entails calculating a Wald ratio for each SNP by dividing the SNP-outcome association by the SNP-exposure association and then estimating the mean of these Wald ratios, weighing each by the inverse of their variances. We used the random-effects model to adjust for heterogeneity among Wald ratios by accounting for over-dispersion in the regression model.

To evaluate whether the IVW estimates comply with the independence and exclusion assumptions, we performed several sensitivity analyses. As a measure of pleiotropy, we assessed heterogeneity among the individual Wald ratios from the initial estimates using I^2^ and Cochran’s Q as well as the intercept taken from the MR-Egger method. We also performed the MR analysis using other methods (e.g., Simple median^15^, weighted median^15^, and MR-Egger^16^) that are more robust than the IVW approach against deviation from the MR assumptions.

Although these methods have less power, estimates from these analyses that are directional discordant from the IVW estimates could be an indication of the presence of pleiotropy. Cook’s distance and the MR Pleiotropy Residual Sum and Outlier^17^ (MR-PRESSO) method were used to identify and remove pleiotropic outliers. SNPs with Cook’s distance > (4/number of SNPs) were tagged as outliers and filtered out due to their disproportionate level of influence on the MR models. MR-PRESSO uses a leave-one-out methodology to detect pleiotropic SNPs and quantifies their distortion in the causal estimate. To ensure the instrumental variables used were the same in our EOS and LOS estimates, we removed non-overlapping SNPs before recalculating the IVW estimate. All statistical analyses were done in R (4.0.3; The R Foundation for Statistical Computing) using the MendelianRandomization (0.9.0) and MR-PRESSO packages (1.0).

Odds ratios were calculated for both EOS and LOS using the final IVW estimate for the casual association of each risk factor with all stroke and toast subtype. To compare the difference of the estimates between early and late onset stroke groups, we performed a t-test, calculated as the difference between the betas divided by the variance of the difference. To account for multiple testing (11 risk factors traits and 6 subgroup analyses), we considered a P-value < 0.00075 (P < 0.05/ (11*6)) to be statistically significant.

## Data Availability

**Individual level data from SiGN, where permitted by participant consent and institutional certification, has been deposited into dbGaP. Summary level GWAS statistics from SiGN and EOSC are available on the Cerebrovascular Disease Knowledge Portal.**

## Results

### Risk Factor vs All Stroke

Figure 2 shows the IVW causal estimates between the stroke risk factors and ischemic stroke for EOS and LOS converted to odds ratios. The odds ratios for continuous risk factors were all based on a per one standard deviation increase except for blood pressure, for which DBP was scaled to a 5 mm Hg increment and SBP to a 10 mm Hg. Because not all odds ratios were scaled the same across traits, caution must be exercised when comparing the relative impact between risk factors and thus we focus on comparisons of each trait between EOS and LOS.

All risk factors except TCHOL were significantly associated with EOS, while only SBP (1.23 OR; 1.15-1.31 95% CI), DBP (1.24; 1.17-1.30), smoking initiation (1.58; 1.09-2.28), T2D unadjusted (1.16; 1.07-1.25), and T2D adjusted for BMI (1.15; 1.07-1.25) were significantly associated with LOS. We further compared the magnitude of causal associations between EOS and LOS. Heterogeneity test indicated that the effect sizes for BMI (odds ratios: 1.52 vs 1.01, Wald’s test p = 1.96E^-08^), DBP (1.53 vs 1.24, p = 1.11E^-09^), and SBP (1.50 vs 1.23, p = 2.28E^-04^) differed significantly between EOS and LOS and nominally for HDL (0.78 vs 0.95, p = 3.37E^-3^) and TG (1.18 vs 1.00, p = 3.51E^-2^).

### Assessment of the MR assumptions

Weighted median, simple median, and MR-Egger were used as alternative causal estimators and their estimates remained stable relative to the IVW estimate (Figure 3). The MR-Egger intercept indicated no evidence for pleiotropy (p > 0.05; **Table S4-5**). There was no strong evidence of heterogeneity among the Wald ratios using I^2^ and the Cochran Q test (I^2^ > 50% and p < 0.05; **Table S6**).

**Figure 3:**
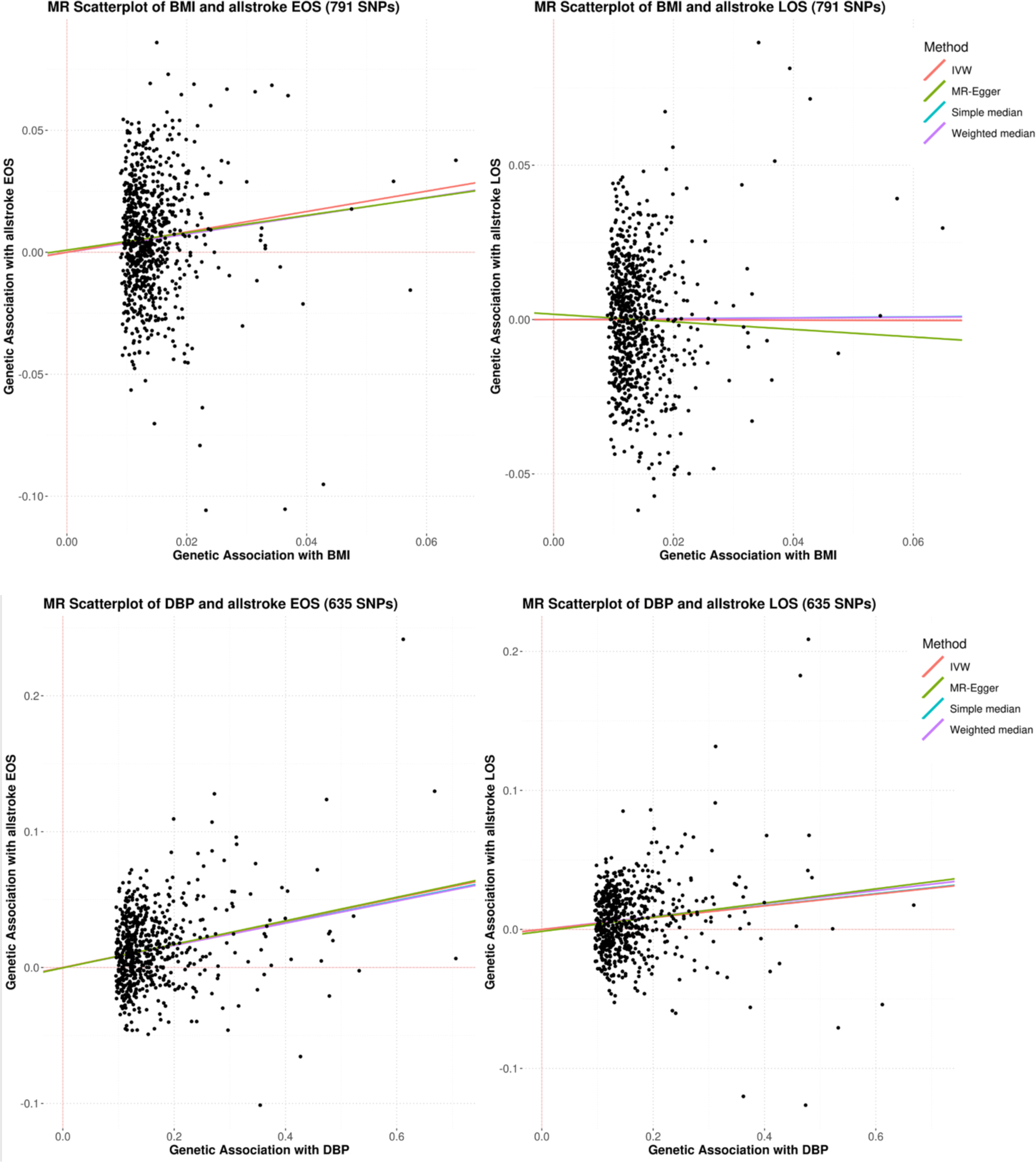
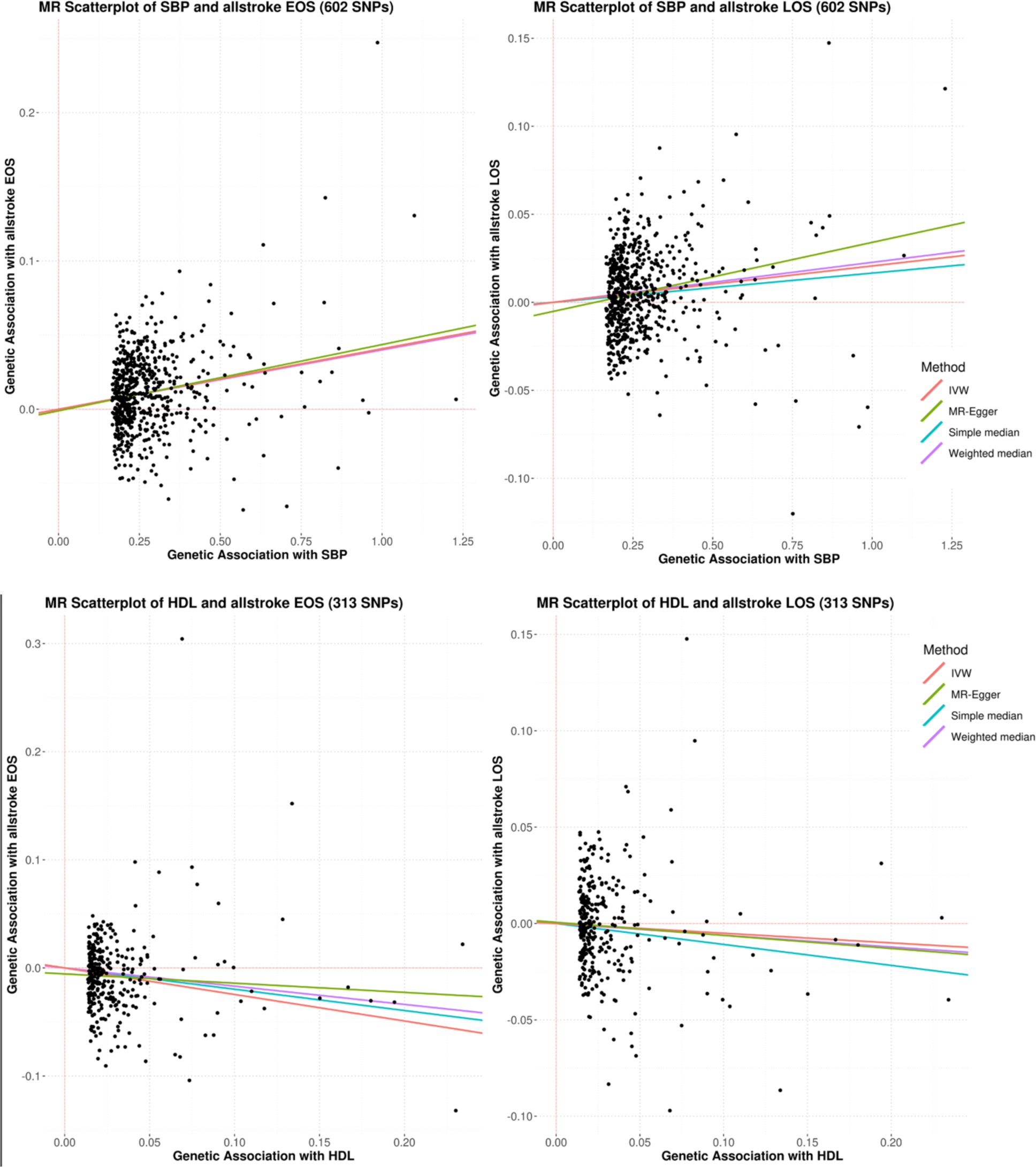
MR Scatterplots and Causal Estimators of BMI, DBP, SBP, and HDL Association with All Stroke EOS and LOS.

### Risk Factor to Toast Subtype

To evaluate whether the stronger associations of BMI, SBP, and DBP with EOS compared to LOS could be attributed to differences in stroke subtypes, we performed subtype-specific MR analyses of these risk factors with the 5 TOAST subtypes. We reasoned that if the EOS vs. LOS differences in risk factor associations were attributable wholly to differences in the distribution of stroke subtypes between EOS and LOS, then there would be no difference in risk factor associations within stroke subtypes. In general, the stronger associations of risk factors observed among those with EOS were preserved within stroke subtypes. For example, BMI was more strongly associated with EOS versus LOS for cardioembolic stroke, large artery atherosclerosis, and small artery occlusion. SBP and DBP were also more strongly associated with EOS compared to LOS within large artery stroke, but only SBP in small arteries (**Figure S1, Table S7**).

## Discussion

The contribution of conventional stroke risk factors to the development of ischemic stroke has been established through prospective epidemiologic studies^18,19^ and causal estimates characterized through MR analyses.^20^ Our study adds to this literature by demonstrating a relatively larger impact of some of these risk factors, namely, higher levels of BMI, and blood pressure, and lower levels of HDL cholesterol, on EOS compared to LOS. Moreover, these patterns generally hold across stroke subtypes, suggesting that differences in the proportions of stroke subtypes across EOS and LOS does not explain the difference in risk factor causal estimates between early and late-onset stroke.

Our results are consistent with prior epidemiologic studies reporting larger effects of some conventional stroke risk factors on early onset stroke. For example, in a case-control study of young ischemic stroke (15-49 years old), Mitchell et al.^21^ found obesity to be significantly associated with an increased risk of ischemic stroke in young adults with an odds ratio of 1.57 (1.28 – 1.94). In other BMI studies, dominated by older onset strokes, odds ratios in the range of 1.02 - 1.30 have been reported.^22^ Similarly, observational studies have shown a stronger dose response for smoking in current versus non-smokers,^23,24^ and hypertension in younger compared to older adults.^25^ In contrast, the association of HDL-cholesterol with ischemic stroke has been reported in at least one study to be stronger in older compared to younger individuals (age <65: OR = 0.76 (0.44-1.32), age 65-74: OR = 0.38 (0.22-0.65), and age ≥75: OR = 0.51 (0.27-0.94)).^26^

The prevalence of many conventional stroke risk factors has steadily risen over the past decades. In a review of the National Health and Examination Survey, Aggarwal et al. found that between 2009 and 2020 the prevalence in hypertension among US adults aged 20-44 years rose from 9.3% to 11.5%, prevalence of diabetes rose from 3.0% to 4.1%, and prevalence of obesity rose from 32.7% to 40.9%.^27^ Concurrent with this rise in stroke risk factors among the young, stroke incidence has increased among younger adults. For example, from 1995 to 2012, US ischemic stroke hospitalization rate increased by 41.5% and 30% for males and females, respectively, aged 35-44 years old.^28^ Among those hospitalized, the prevalence of traditional risk factors was nearly doubled, where one in three men had three to five risk factors. Thus, the increased prevalence of stroke risk factors among the young, combined with the greater impact they have on younger adults, may partly explain the rising incidence of ischemic stroke in this age group.

A major strength of our study is the use of Mendelian randomization to estimate the causal effects of conventional stroke risk factors on early and late onset stroke cases. The genetic instruments were based on summary statistics obtained from large genome-wide association studies. We further assessed the effects of the risk factor instruments on well characterized early and late onset stroke cases and controls assembled through large consortia with thorough endpoint validations.

Like many studies, a major limitation of our study is its restriction to individuals of European ancestry, due primarily to the relatively small contribution of non-European samples to existing genome-wide association studies of stroke risk factors and stroke. Future studies involving non-European samples are urgently needed.^29^

In summary, this study uses large, publicly available GWAS summary statistics to conduct MR analyses to estimate causal effects of conventional stroke risk factors on early and late onset stroke. We found that genetically predicted, higher levels of BMI, DBP, and SBP and lower levels of HDL-C were more strongly associated with risk of EOS compared to LOS. These results highlight the need to address the rising prevalence of conventional risk factors, particularly BMI and blood pressure among young adults.

